# Analysis of the Clinical Incidence and Correlation between Colorectal Cancer and Microorganisms

**DOI:** 10.1101/2023.06.14.23291347

**Authors:** Kalia Koutouvalis, Pablo A Bejarano

**Author notes:** Corresponding author: Kalia Koutouvalis.

## Abstract

In this single institution retrospective medical record review, patients diagnosed with colorectal cancer from the years 2018-2022 were evaluated to distinguish an associative linear relationship between diagnosed colorectal cancer and a positive result for the presence of a microorganism. Based on the clinical incidence of this occurrence, it was observed patients with tumors in the left side of the colon had a higher incidence of a positive test result with a dominant microorganism. Species evaluation within this cohort found similarity to microorganisms identified as colorectal cancer biomarkers. These findings support clinical relevance and warrant further consideration for prospective study regarding microorganism involvement in colorectal cancer.

## Introduction

Colorectal cancer (CRC) is the fourth most prevalent cancer in the United States. In 2023 it is estimated approximately 150,000 individuals will be diagnosed with CRC (Siegel,Wagle et al. 2023). Identifying and managing risk factors related to CRC has reduced the disease burden amongst diagnosed patients. According to the Centers for Disease Control and Prevention, the following are the most prominent risk factors and contributors to the development of CRC. Age, inflammatory bowel diseases, like ulcerative colitis (UC) and Crohn’s disease (CD), genetic or epigenetic predisposition and lifestyle factors, including tobacco use, alcohol consumption, a high-fat low fiber diet and insufficient physical activity. The mechanism established in the development of CRC is most commonly the alteration of normal colonic epithelial cells to carcinoma through the adenoma-carcinoma sequence (Grady and Carethers 2008). This sequence typically follows three routes of tumorigenesis including chromosomal instability (CIN), microsatellite instability (MSI) and the serrated pathway. These routes are characterized by DNA manipulation and mutations leading to the progression of malignant neoplasms (Molavi, et al .2018, Currais, Rosa et al. 2022, Ryan, Sheahan et al. 2017).

Due to the specific pathology of these neoplastic lesions, the microbiota that tend to colonize within the intestinal crypts have been associated with potential mechanisms of inducing and/or mediating tumorigenesis (Jin, Shang et al. 2021, Kang, Zhang et al. 2021, Keku et al. 2015). One possible mechanism related to microbial associated tumorigenesis is through the host’s immune system. Signaling pathways associated with regulation of host immune responses and tumor suppression in early oncogenesis have been implicated with various members of the microbiome (Bauché and Marie 2017, Daniel, Ball et al. 2017, Pang, Tang et al. 2018). There are a variety of microbiota suggested to integrate into the tumor microenvironment promoting tumorigenesis that have now been distinguished as biomarkers for CRC specific microbiome configurations. These microorganisms include, *Fusobacterium nucleatum*, *Bacteroides fragilis*, *Streptococcus, Prevotella*, *Clostridium difficile*, *Clostridium symbiosum, Escherichia coli, and Enterococcus faecalis,* including indication of their respective metabolomic signatures (Sillo, Beggs et al. 2023, Keku et al. 2015, Lin et al. 2022, Pop et al. 2020, Liu N.N. et al 2022).

Information on the mechanisms in which these microbial species mediate tumorigenesis is limited, however trends established between the microbiome and CRC tumors may provide further insight. For instance, certain microbial species have been linked to specific tumor types and locations within the gastrointestinal tract in patients with CRC (Jin, Shang et al. 2021, Liu, N.N. et al. 2022, Dominik et al. 2020). Based on trends analyzed within pre-cancerous lesions, a CRC specific microbial phenotype has been identified, where combining screening methods with metagenomic sequencing ultimately has displayed a less invasive functionally efficient screening approach (Kværner et al. 2021, Lee W.J.J. et al., 2023).

Interaction between microbiota and the tumor microenvironment may be more complex than any single correlation between changes within the microbiome and carcinoma precursors, which may involve a variety of factors, including age, diet, microbiome configuration, genes associated with oncogenesis and external environment working in syncytium (Watson et al. 2023, Pang et al. 2018). A prevalent confounding factor in the gut microenvironment of CRC is inflammation, unregulated microbial interactions and the inability to modulate immune anti-carcinogenic pathways (Elinav et al. 2013, Kim and Lee 2022, Wang and Li 2022, Lamaudiere et al. 2023). The appearance of commensal or pathogenic microorganisms interacting with the gut microenvironment contributing to gastrointestinal dysbiosis suggests these findings may manifest clinically. This report aims to investigate the clinical incidence of a dominant microorganism, whether causing overt disease or not, accompanying a formal diagnosis of CRC to consider the frequency and correlation of microorganism involvement and potential use of the microbiome as a CRC screening method in this specific clinical setting.

## Methods

This medical record review was approved by Cleveland Clinic’s IRB and all information present followed the submitted research protocol and data collection sheet. A population of individuals who have been treated in Cleveland Clinic Weston Florida for their diagnosed CRC between February 1^st^, 2018, and February 1^st^, 2023, was compiled using Cleveland Clinic’s eResearch databank. The population pool ensured the individuals considered were over the age of 18 and under the age of 90 and had no actively treated autoimmune disorders during the period of interest. This population was then filtered for whether there was an order or test placed for the microbiology laboratory within the patient’s medical record. The focus of this search included tests with the purpose of ruling out or identifying a microorganism during the period of interest. This was further filtered to ensure patients have not taken any course of antibiotics, excluding topical antibiotics, within the two-month time frame prior to the serological, histological, or laboratory test being collected and sent for testing. Information presented within the patients’ medical record included testing facilities outside Cleveland Clinic if pertinent to the inclusion criteria related to antibiotic administration. Information on the CRC including histological type, tumor size, location, and differentiation as well as the type of order/test and corresponding result were entered into Cleveland Clinic’s Redcap database. The Redcap database allowed organization of CRC cases in a format where all cases were entered in a randomized order and assigned a new case number only relevant to this study so that no one outside the research group can identify the study subjects.

### Data Analysis

Study data were collected and managed using REDCap electronic data capture tools hosted at Cleveland Clinic (PA Harris et al. 2009, PA Harris et al. 2019). REDCap (Research Electronic Data Capture) is a secure, web-based software platform designed to support data capture for research studies, providing 1) an intuitive interface for validated data capture; 2) audit trails for tracking data manipulation and export procedures; 3) automated export procedures for seamless data downloads to common statistical packages; and 4) procedures for data integration and interoperability with external sources. Data was analyzed using a linear model on Cleveland Clinic provided statistical software.

## Results

### Population Characteristics

The study cohort included 241 total patients that comprised of 214 adenocarcinomas, 7 mucinous adenocarcinomas, 2 medullary carcinomas, 4 signet-ring cell carcinomas, 11 squamous cell carcinomas and 2 neuroendocrine carcinomas, with one adenocarcinoma having neuroendocrine differentiation noted (Table 1). The population combined all individuals who were diagnosed with CRC that also had orders/tests placed with the purpose of ruling out and/or identifying microorganisms or being sent to the microbiology laboratory (Figure 1). Among the patients who fit the criteria, 101 had microbiology related orders and tests solely relating to the SARS-COV-2 virus. These cases will be noted (Supplemental Table 1), however they are not the focus of this investigation. Out of the remaining 140 cases that were not related to the SARS-COV-2 virus, 50.6% (n=71) had a dominant microorganism present within a test result.

**Figure 1:**
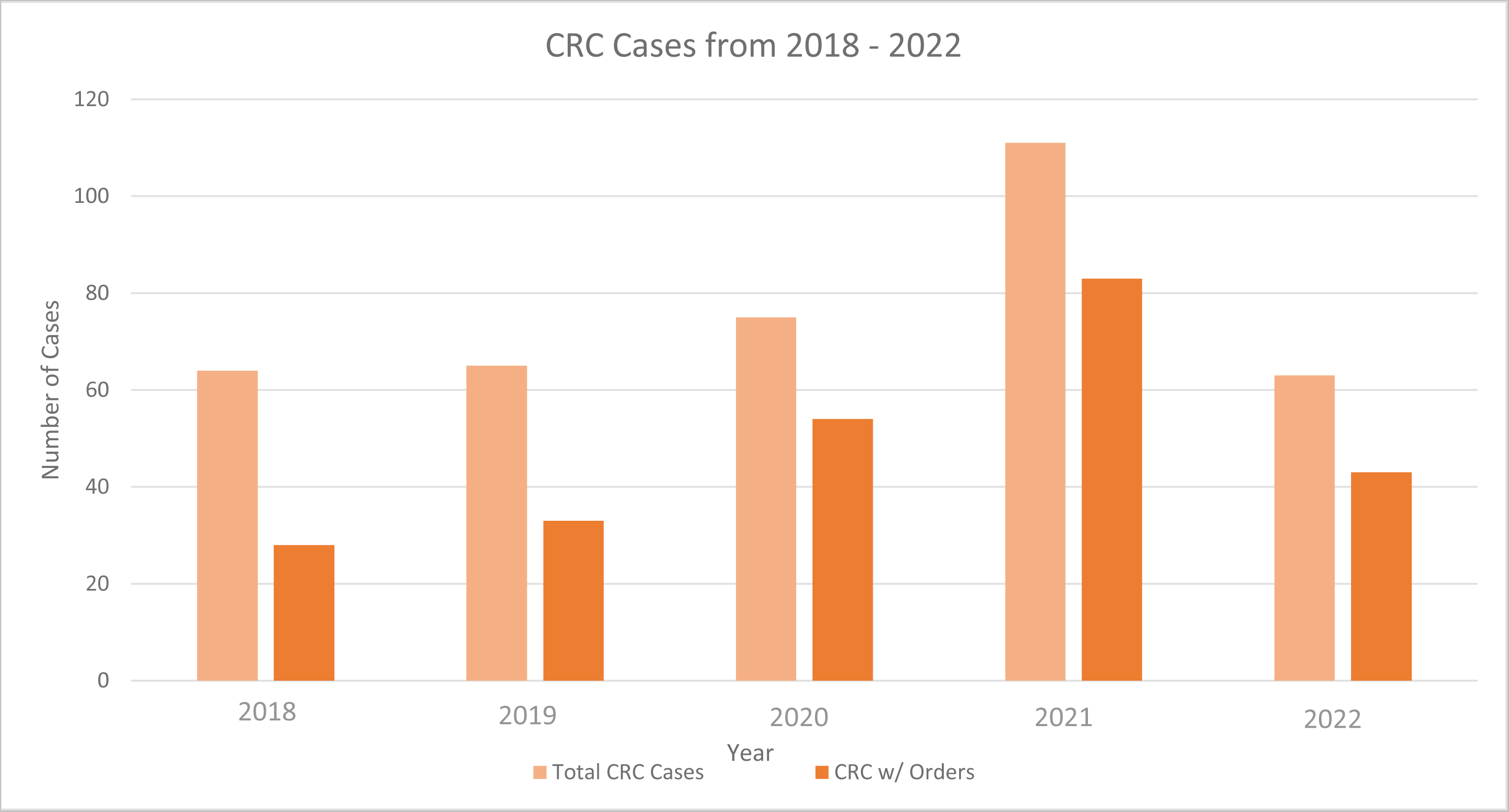
Total Cases of CRC compared to Cases of CRC with Orders/Tests placed for microbiology A comparison of the total CRC cases diagnosed in Cleveland Clinic Florida’s Weston campus compared to CRC cases that have a corresponding test/order placed for ruling out or identifying a microorganism.

**Table 1:**
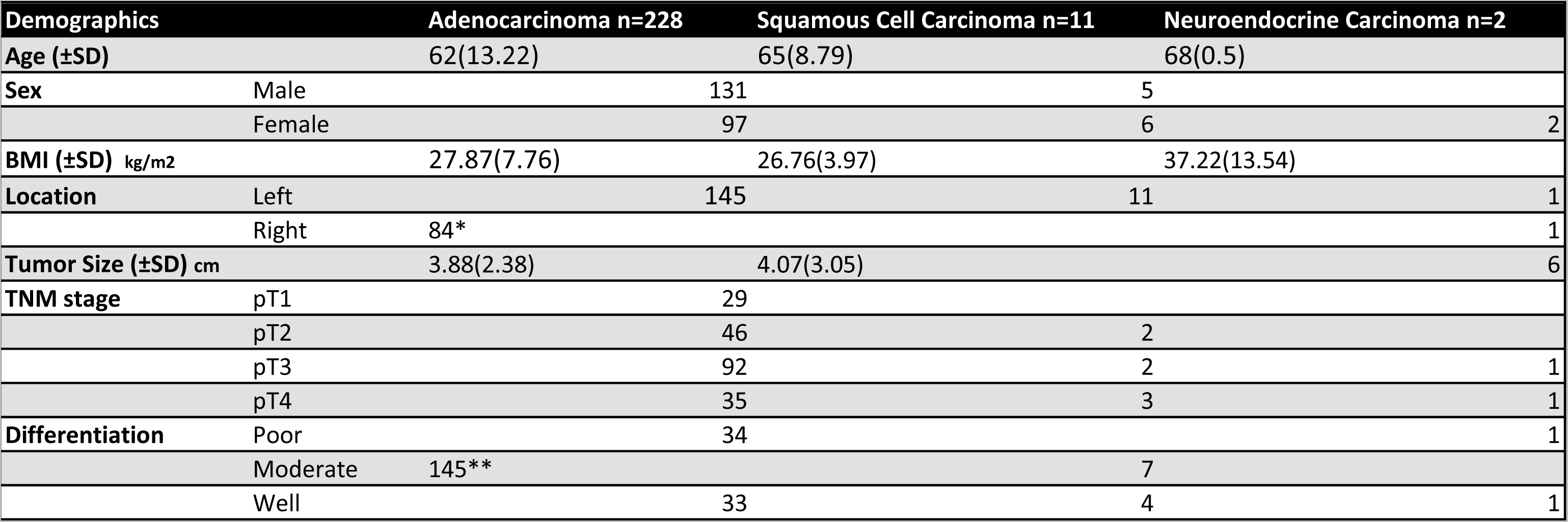
Population Demographic Coordinated by CRC Type * One case had two tumors, each respectively located in the left and right colon ** The instance in which a case was classified as “moderate to poorly differentiated” is categorized under “Moderate” in this table

### Linear Model Analysis

To determine a relationship between CRC cases and orders relating to microbiology a linear regression was run on the n= 378 cases of diagnosed CRC in Cleveland Clinic’s Weston Campus and the n=241 cases fitting the criteria of this investigation. It was found that over the five-year period considered, CRC patients who have had orders or tests placed with the intent to identify or rule out a microorganism compared to the total number of patients diagnosed with CRC, were statistically significant in relation to each other (p<0.001) at the 95% confidence interval (Figure 2A, Supplemental Table 2). Interpreting the residual plot for the total cases of CRC versus CRC cases with orders for microbiology displays data points not fitted around zero (Figure 2B). This indicates the possibility of a relevant variable not being considered within this model. For the scope of this report, the variables within the limit of discussion and data collection are those which are included. The relationship between the n=140 cases with microbiology related orders and n=71 cases which had a dominant microorganism present was also considered. The results of this linear regression analysis determined cases that had a dominant microorganism were significantly related (P<0.001) to the tests that had microbiology orders placed not relating to SARS-COV-2. Although this statistical significance is present between the two variables, the regression model explains 37% of variability (Figure 3A & Supplemental Table 3) and suggests more relevant variables in addition to a better fitting model need to be considered to provide meaningful analysis (Figure 3B). This could include considering the (n=101) SARS-COV-2 related cases that were omitted in this analysis, to account for the observations not represented in-between the data points present.

**Figure 2:**
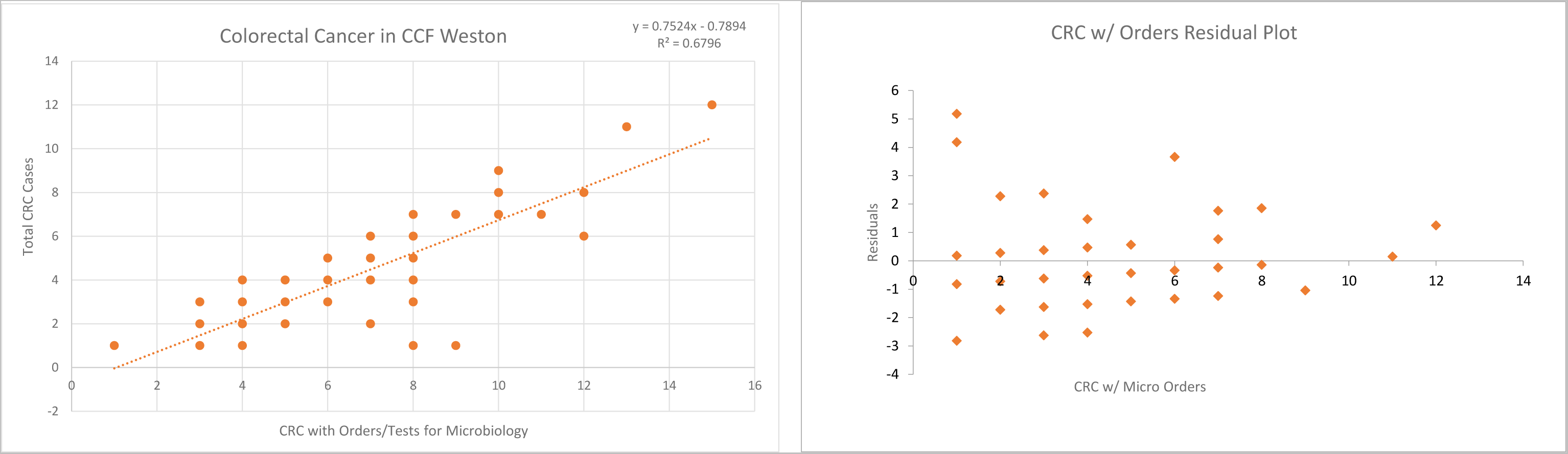
Linear Regression of Total Cases The regression analysis based on total CRC cases seen in the Cleveland Clinic Weston campus from 2018-2022 (A) the linear regression scatter plot supporting 68% of all variability in the data set is explained by the regression model and (B) the data presented on a residual plot.

**Figure 3:**
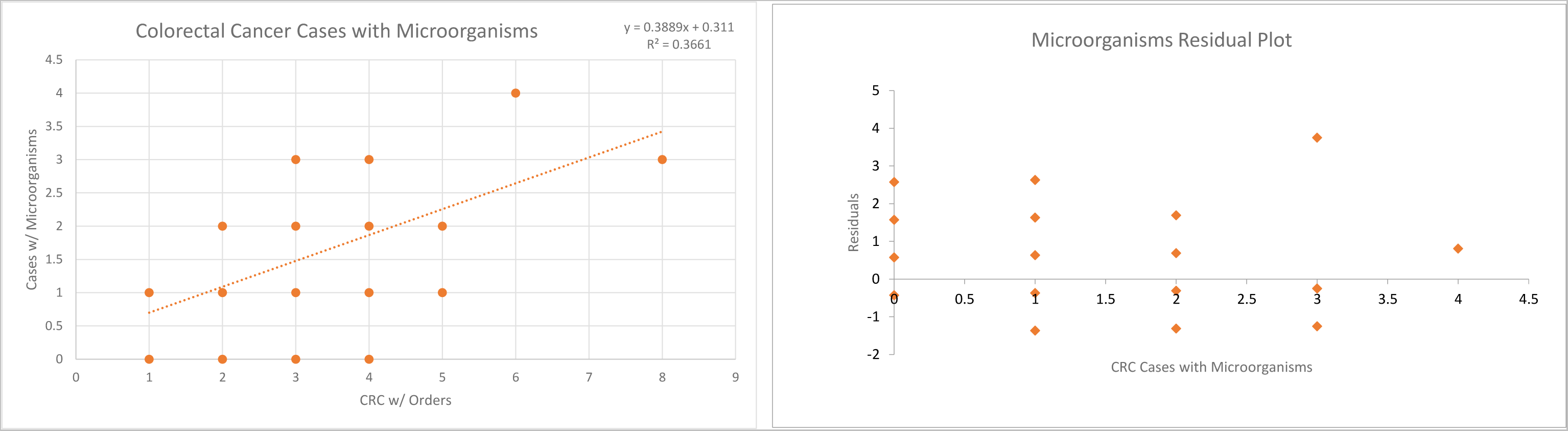
Linear Regression of Cases with Microbiology Association Regression analysis representative of the cases of CRC that have microbiology related orders correlated to cases presenting with organisms (A) the linear regression representing 37% of variability within the model and (B) the residual plot presenting with a similar trend to the regression plot for CRC cases with orders placed where the residuals are not plotted around zero.

Moreover, it can be speculated these residuals may be due to an increase and high importance of SARS-COV-2 related microbiology orders being placed during the year 2020 and therefore a decrease in the performance rate of the remaining tests. However, the data being presented cannot make definitive confirmation that SARS-COV-2 impacted patients who were diagnosed with CRC having a positive test result accompanying an identified microorganism (Eklöv K. et al. 2022, Blondeau J.M. 2020).

### Microbiology Tests/Orders and Dominant Microorganisms

Amongst the (n=140) CRC cases that fit under the criteria of having orders and tests placed with the purpose of identifying or ruling out a microorganism or being sent to the microbiology laboratory, there were a total of 419 orders/tests placed. The most frequent and abundant tests were urine and blood cultures followed by H&E/IHC stains ordered on surgical specimens with the purpose of ruling out microorganisms within the tissue (Figure 4). The largest amount of gathered data related to tests/orders in respect to CRC cases was observed under the adenocarcinoma diagnostic category. Left colon adenocarcinomas had approximately 50% more urine cultures ordered than in right colon adenocarcinomas. A similar trend was seen with blood cultures. There was a greater number of H&E/IHC stains ordered for right colon adenocarcinomas. In addition, right colon adenocarcinomas displayed a greater incidence for respiratory/pulmonary related tests (Table 2). These tests produced a total of 109 positive results for microorganisms present within the collected samples along with their respective frequency of result and identification (Figure 5). The tests being discussed do not include any SARS-COV-2 related tests despite their presence within the patient’s medical record during the period of interest.

**Figure 4:**
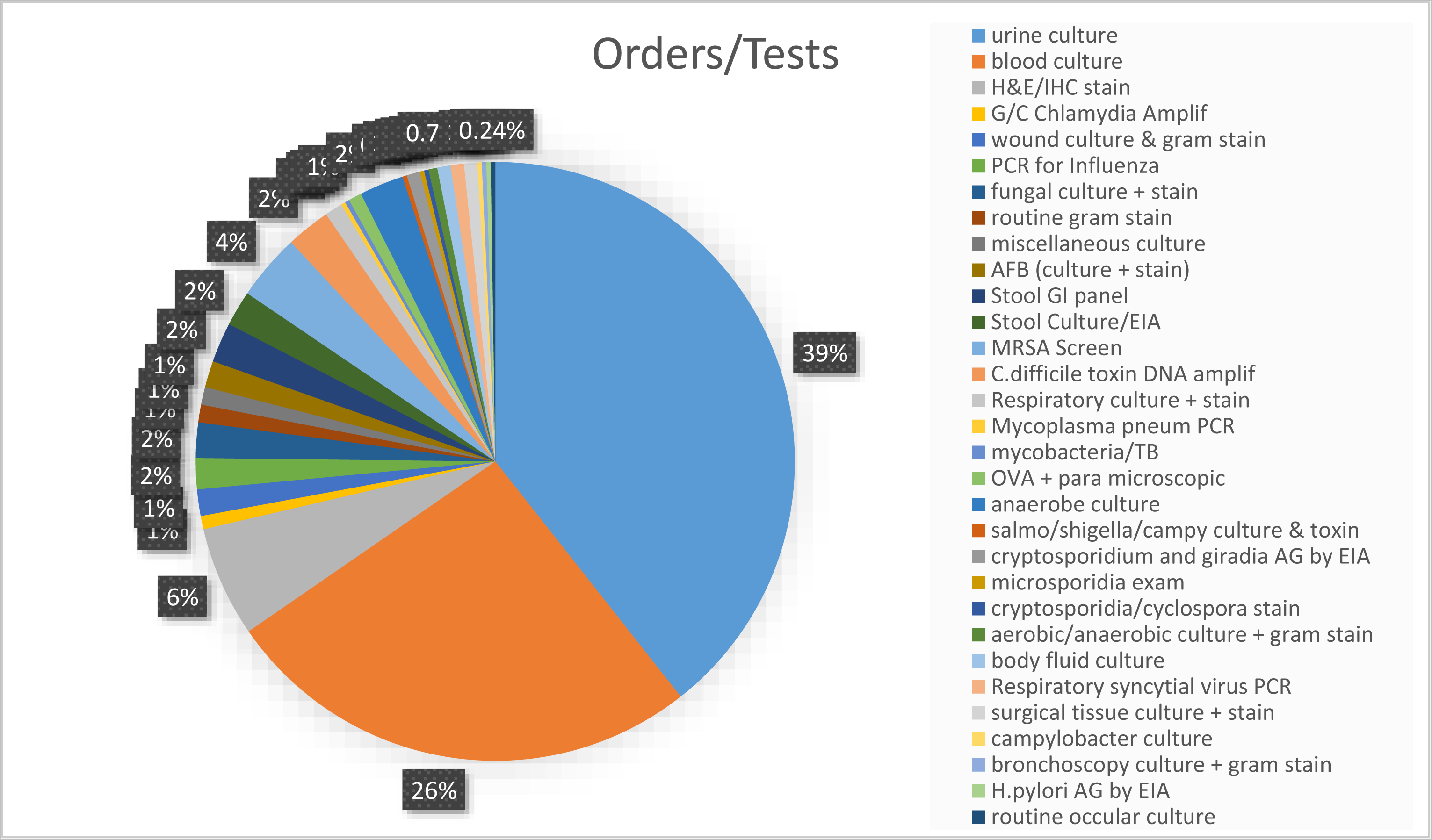
Total Tests Ordered and Listed Corresponding to CRC cases A display of the proportion of tests/orders placed corresponding to the institutional nomenclature of the test/order conducted.

**Figure 5:**
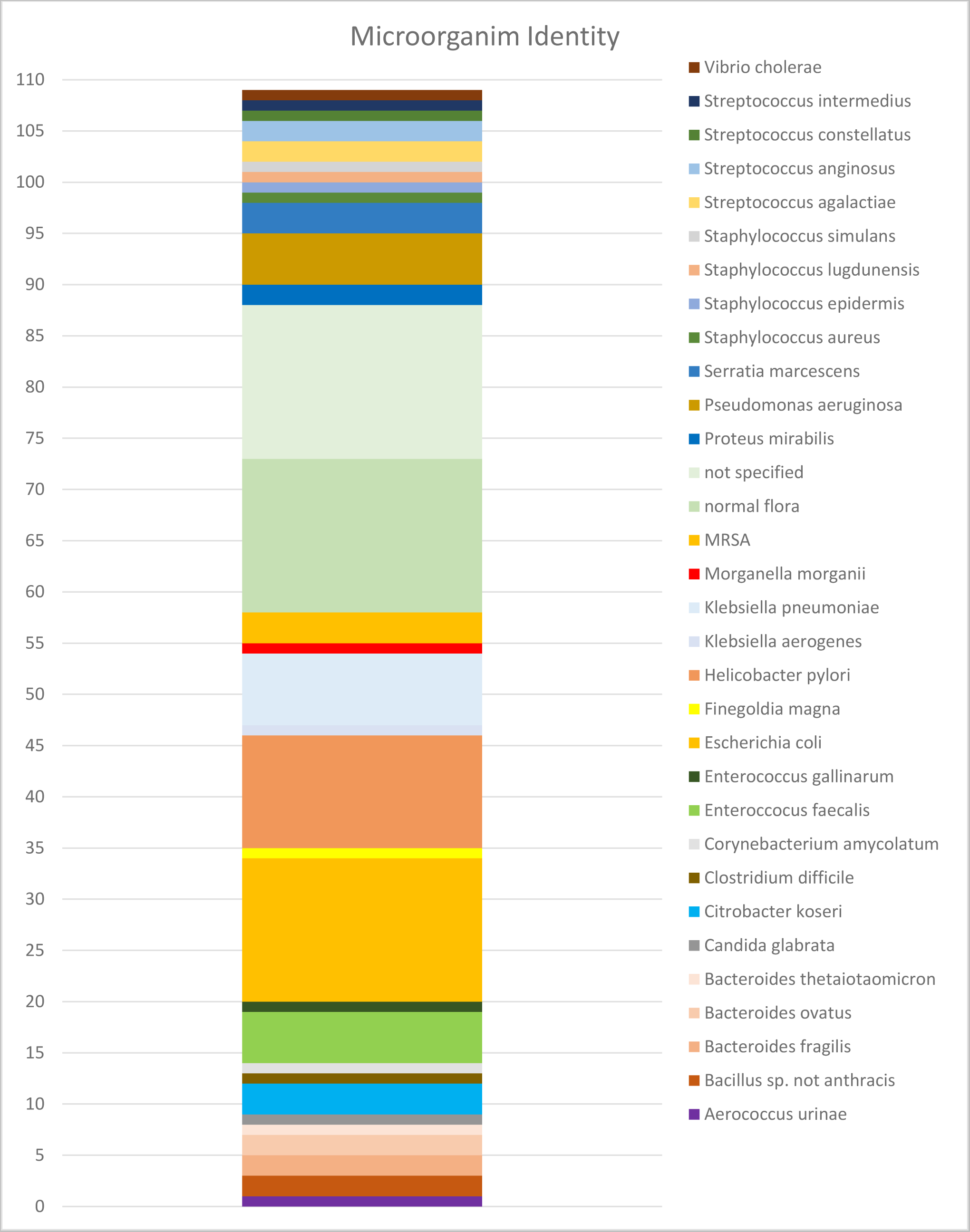
Microorganism Identities & Abundance Based on Degree of Identification Distribution of microorganism classification based on identification within a positive test result.

**Table 2:**
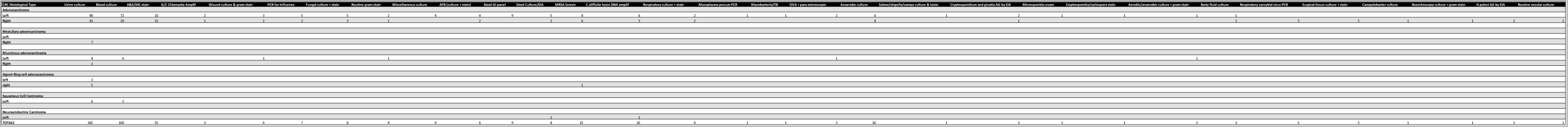
Microbiology Related Orders/Tests in Respect to CRC Characteristics

One prominent category discovered during data collection was observed when tests displayed a positive result, however there was no distinctive identification or further notation of this result. These instances fell under the category of positive test result with unspecified organism further referred to as ‘not specified.’ Another prominent category encountered was, a result although noted as a positive for the presence of microorganisms, was not a cause for concern due to microbiota present consisting of the normal flora found within the tested specimen type. These categories made up a total of 30 outcomes within the 109 positive results. These two categories will not be further analyzed as their results are nonspecific and do not provide insight into trends between dominant microorganisms and CRC.

The microorganisms with the highest prevalence within CRC cases were *Escherichia coli*, *Helicobacter pylori*, *Klebsiella pneumoniae*, *Enterococcus faecalis*, *Pseudomonas aeruginosa*, *Staphylococcus*, *Streptococcus* and *Bacteroides*. Comparing these results to the cases that had orders/tests placed for microbiology, 65 were adenocarcinomas, 3 were mucinous adenocarcinomas, 2 signet-ring cell carcinomas, and 2 squamous cell carcinomas, one of the adenocarcinomas had neuroendocrine differentiation noted. The 2 previously mentioned neuroendocrine carcinomas did not have a dominant microorganism present. Left sided CRCs were favored in respect to presence of a dominant microorganism in comparison to the right side (Figure 6A). Within the cases of left sided CRCs *E. coli* was the most ubiquitous. In addition, although at a lower prevalence, both *Enterococcus* species appeared in test results of left sided CRC’s. The majority of the microorganisms that occur at a higher prevalence are correlated with cases of left side CRC’s. This is not true for *H. pylori*, *Staphylococcus*, *Streptococcus* and *Bacteroides* that are split evenly in both right and left sided CRC’s (Figure 6B).

**Figure 6:**
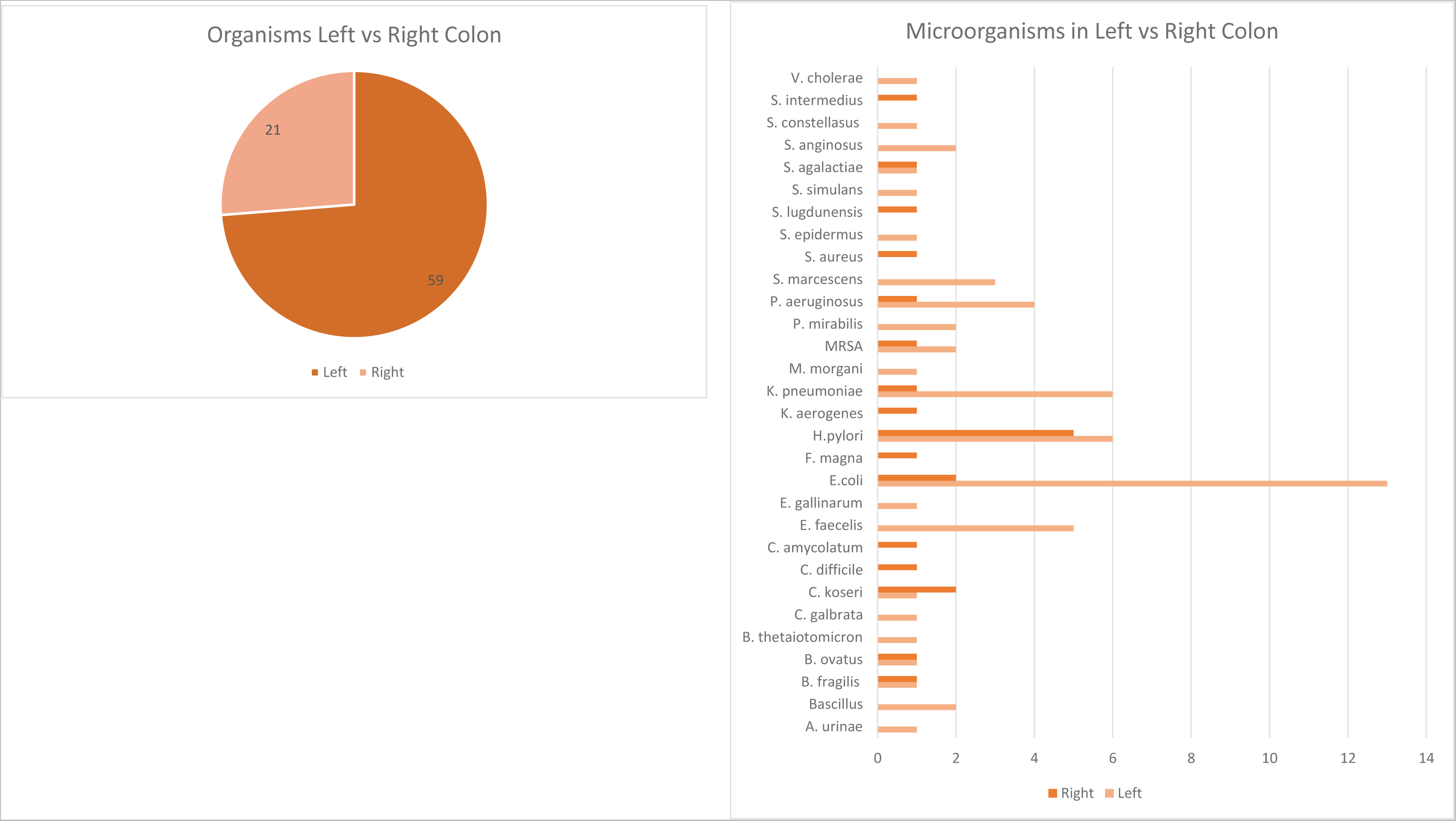
Microorganism correlated to CRC location within GI tract A representation of microorganism corresponding to location of CRC within the GI tract

These 109 test results indicating presence of a dominant microorganism were shared amongst a group of 71 patients with a vast overlap occurring in a few of these patients. This is seen particularly in three CRC cases that have the most microbial presence within their reports. They comprise of the following organisms; *A. urinae*, *B. fragilis*, *B. ovatus*, *B. thetaiotaomicron*, *C. koseri*, *E. faecalis*, *E. coli*, *P. aeruginosa*, *P. mirabilis*, *S. simulans*, and *S. anginosus*. One case, a moderately differentiated mucinous adenocarcinoma of the left colon diagnosed in a male over the age of 80 with a BMI greater than 30, had positive test results for 7 of these listed organisms. In comparison, the remainder of the positive test results were represented by a sole occurrence under one patient. This was seen with *E. gallinarum*, *K. pneumoniae* and some *Bacteroides*. The sole presence of *Corynebacterium amycolatum*, *Staphylococcus lugdunensis* and *Finegoldia magna* occurred in the same CRC case of a moderately differentiated adenocarcinoma in the right colon female with a BMI over 50 that also had a result positive for *Pseudomonas aeruginosa*. The sole presence of *Klebsiella aerogenes* was in an ocular sample, further correlation to CRC is not being considered for this organism. *Escherichia coli* was predominantly present in males with moderately differentiated adenocarcinoma in the left colon (Table 3). Presence of *Helicobacter pylori* did not follow any trends based on cancer characteristics, sex, age or BMI.

**Table 3:**
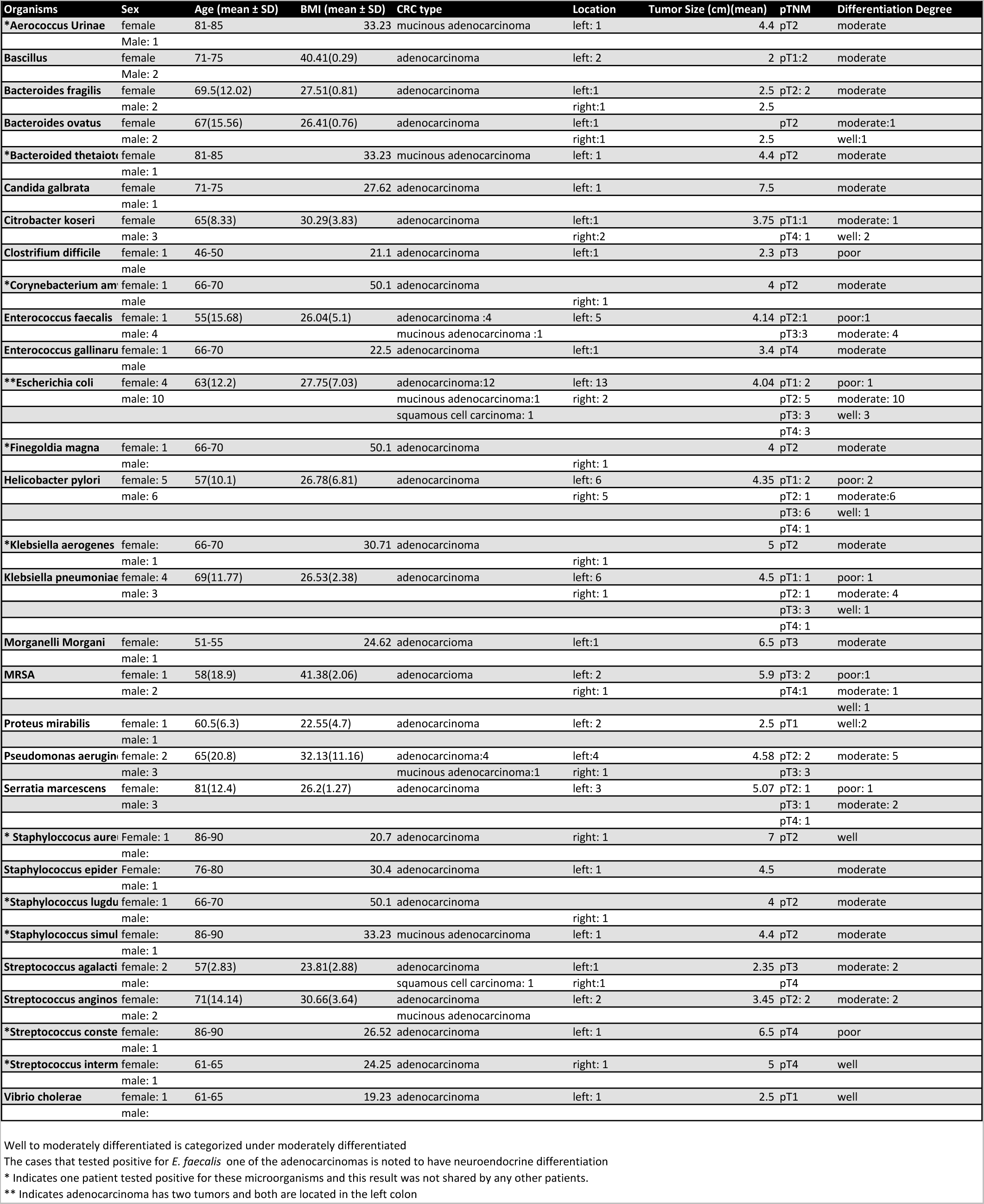
Microorganisms Related to Patient Demographics and CRC Characteristics Well to Moderately Differentiated is categorized under moderately differentiated. Out of the cases that tested positive for E. faecalis, one of the adenocarcinomas are noted to have neuroendocrine differentiation. * Indicates one patient tested positive for these microorganisms and this result was not shared by any other patients. ** Indicates adenocarcinoma has two tumors and both are located in the left colon.

### Demographics and Social Determinants of Patients with Dominant Microorganisms

The 71 patients that had a positive test result with a dominant microorganism had an average age at time of diagnosis of 63 years (SD 13.9) and an average BMI of 29.08 kg/m^2^(SD 11.3). 39 were male and 32 were female. 36 had co-occurrent gastrointestinal diseases that included 3 ulcerative colitis, 1 Crohn’s disease, 10 Type II diabetes mellitus (T2D), 19 Gastroesophageal Reflux Disease (GERD), 5 that had both T2D and GERD and 2 other cooccurring GI disorders. These trends do not correlate with microorganism presence as the incidence of cooccurring GI disparities among the study population remained the same and concur with inflammatory bowel disorders classified as risk factors for the development of CRC. The additional factors discussed as risks for development of CRC involve social determinants of health including tobacco use, alcohol consumption, insufficient physical activity, diet and psychological adversity including depression and stress. This institution documents these factors through self-reported surveys and questionnaires at time of hospital admission.

During data collection, information regarding status of these social determinants was included to investigate whether trends can be observed from the total study population compared to those cases with a dominant microorganism. 84% of the study population had information present regarding tobacco use. This institution records tobacco use in three categories. Low risk, indicating the patient has never used a tobacco product, medium risk indicating the patient is a former user of tobacco products and high risk indicating the patient currently uses tobacco products. A comparison of tobacco usage yields no distinct trends regarding this risk factor in relation to having a positive result of a microorganism. Although not determinative of any result or analysis it is interesting to note the patient that tested positive for the greatest number of microorganisms was classified under low risk of tobacco use. Additional consideration of social determinants including alcohol consumption, physical activity, depression, stress and food security cannot be included as sufficient analysis between the total study population and those with dominant microorganisms is not accurately reflective based on the volume of unreported values for these social determinants at the time of CRC diagnosis.

## Discussion

Colorectal Cancer cases in Cleveland Clinic’s Weston Florida campus were compiled to determine whether there was a correlation between clinical presentation of a dominant microorganism within the patients’ medical record and the formal diagnosis of CRC during a 5-year period. There was statistical significance observed in both of the linear regressions. Although the significance suggested there was a linear correlation between both instances, it appears a linear model was unable to account for all relevant variables necessary within this statistical performance. This may include variables revolving around the SARS-COV-2 virus and the prevalence of tests being ordered in all patients admitted in Cleveland Clinic’s Weston campus compared to patients with CRC. This model can be altered to emulate linear models associated with progression of cancer alongside tests placed to determine microorganism involvement, however the information necessary is beyond the scope of this medical record review (Sung, S.Y et al. 2022, Vuik F.E et al. 2019).

Evaluating the tests placed during the period of CRC diagnosis suggested left colon adenocarcinomas had the greatest prevalence of urine and blood cultures ordered when compared to right CRC. Left colon adenocarcinomas were also at a greater representation in tests that had a positive result for a microorganism. To strengthen this observation, this study’s population pool was compared to a previously established study population of CRC cancers in Cleveland Clinic from 2000 to 2008 by Chouhan and colleagues (Chouhan et al. 2019). The similarities between the demographics and cancer characteristics of these two populations indicated the trend of left sided CRC cases was based on the positive result of a dominant microorganism rather than population characteristics. This could indicate a correlation between left sided CRC tumors and a positive result indicating the presence of a microorganism potentially due to the GI microenvironment, composition and structure of the microbiome, tumor type and interactions with host immune system in this area of the colon (Baran et al. 2018, Zhong et al. 2020).

Although tests provided in our reference laboratory do not focus on microbiome composition, species identified as CRC biomarkers were observed at a high prevalence within this population pool (Sillo, Beggs et al. 2023, Keku et al. 2015, Lin et al. 2022, Pop et al. 2020, Liu N.N. et al 2022). This includes *Escherichia coli*, *Enterococcus faecalis*, and *Streptococcus*. *Helicobacter pylori*, which was among the microorganisms observed at a higher prevalence, has also been associated with CRC and adenomatous polyps (Teimoorian et al. 2018). Even so, it is essential to consider due to CRC’s having high rates of inflammation and CRC specific microbiome phenotypes, it is possible these microbiotas are enriched based on the environmental conditions rather than being prognostic upstream factors (Watson et al., 2023). At our institution Fecal Occult Blood Tests (FOBT) are offered as a screening method. It can be considered to utilize this screening method in addition to metagenomic sequencing to integrate a less invasive screening method within the CRC population (Khannous-Lleiffe et al. 2023, Zwezerijnen-Jiwa et al. 2023, Zhou et al. 2022).

## Conclusion

Although presence of a dominant microorganism within patients that have CRC does not explicitly display correlation, our findings suggest clinical relevance and potential for prospective study. From a clinical standpoint the incidence of a dominant microorganism more likely to occur in left sided CRC may be useful in the management of such infections (Braumüller et al. 2023, Tripathy et al. 2021). Improvement in screening methods and the use of the microbiome as a diagnostic marker display evident potential.

## Limitations

The data collected regarding cooccurrence of tests/orders placed for microbiology testing was not considered for linearity; there is no analysis of up or downstream effects based on the cooccurrence of both variables. The collection of the study population was gathered from an eResearch search engine rather than a Cancer Biobank and therefore may not be fully representative of the CRC cases diagnosed in this singular institution. Individual factors in a patient’s medical history that may have contributed to certain pathology or morbidity that were not within the criteria of this medical record review have not been accounted for. The findings were limited to the available services within the microbiology department including inability to further classify organisms that fell under the ‘not specified’ category along with result limitation to culturable microorganisms. The stool cultures and GI panel tests did not display the presence of a microorganism but rather identification of enterotoxins related to common food-borne or opportunistic pathobionts. The pandemic altered the allocation of resources towards treating the SARS-COV-2 pandemic displayed a greater influx of SARS-COV-2 related microbiology orders. Even so, this population pool did not display an overall decrease in diagnosis of CRC during the pandemic when compared to the years prior as originally anticipated (Freund & Wexner, 2022).

## Supporting information

Supplemental Tale 3

## Data Availability

All data produced in the present work are contained in the manuscript

## Notes

### Competing Interest Statement

The authors have declared no competing interest.

### Funding Statement

This study did not receive any funding

### Author Declarations

The IRB of Cleveland Clinic Florida gave ethical approval this minimal risk research using/involving secondary research for which consent is not required and the research involves only information collection and analysis involving the investigator's use of patient health identifiers when that use is regulated by HIPAA for the purposes of health care operations, research, or public health activities and purposes.

