## Supplemental Table 2 for "Analysis of the Clinical Incidence and Correlation between Colorectal Cancer and Microorganisms"

SUMMARY OUTPUT

| *Regression Statistics* | |
|---|---|
| Multiple R | 0.824359 |
| R Square | 0.679567 |
| Adjusted R Square | 0.673521 |
| Standard Error | 1.649641 |
| Observations | 55 |

ANOVA

| | *df* | *SS* | *MS* | *F* | *Significance F* |
|---|---|---|---|---|---|
| Regression | 1 | 305.879407 | 305.8794 | 112.4013 | 1.04716E-14 |
| Residual | 53 | 144.2296839 | 2.721315 | | |
| Total | 54 | 450.1090909 | | | |

| | *Coefficients* | *Standard Error* | *t Stat* | *P-value* | *Lower 95%* | *Upper 95%* | *ower 95.0%* | *pper 95.0%* |
|---|---|---|---|---|---|---|---|---|
| Intercept | 2.915196 | 0.434532994 | 6.708802 | 1.33E-08 | 2.043633076 | 3.786759 | 2.043633 | 3.786759 |
| X Variable 1 | 0.903171 | 0.08518915 | 10.60195 | 1.05E-14 | 0.732303266 | 1.074039 | 0.732303 | 1.074039 |

RESIDUAL OUTPUT

| *Observation* | *Predicted Y* | *Residuals* | *dard Residuals* |
|---|---|---|---|
| 1 | 6.52788 | 1.47211986 | 0.900768 |
| 2 | 5.624709 | 0.375290923 | 0.229635 |
| 3 | 4.721538 | -1.721538014 | -1.05338 |
| 4 | 6.52788 | 0.47211986 | 0.288883 |
| 5 | 5.624709 | 0.375290923 | 0.229635 |
| 6 | 3.818367 | -0.818366951 | -0.50075 |
| 7 | 4.721538 | 2.278461986 | 1.394156 |
| 8 | 5.624709 | 2.375290923 | 1.453404 |
| 9 | 4.721538 | 2.278461986 | 1.394156 |
| 10 | 5.624709 | -0.624709077 | -0.38225 |
| 11 | 3.818367 | 0.181633049 | 0.111138 |
| 12 | 4.721538 | -1.721538014 | -1.05338 |
| 13 | 3.818367 | 4.181633049 | 2.558677 |
| 14 | 6.52788 | 0.47211986 | 0.288883 |
| 15 | 3.818367 | 5.181633049 | 3.170562 |
| 16 | 4.721538 | -0.721538014 | -0.4415 |
| 17 | 3.818367 | -0.818366951 | -0.50075 |
| 18 | 6.52788 | -2.52788014 | -1.54677 |
| 19 | 6.52788 | -1.52788014 | -0.93489 |
| 20 | 3.818367 | 0.181633049 | 0.111138 |
| 21 | 6.52788 | 0.47211986 | 0.288883 |
| 22 | 8.334222 | -1.334222265 | -0.81639 |
| 23 | 5.624709 | -1.624709077 | -0.99413 |
| 24 | 4.721538 | 0.278461986 | 0.170387 |
| 25 | 6.52788 | -1.52788014 | -0.93489 |
| 26 | 6.52788 | 0.47211986 | 0.288883 |
| 27 | 5.624709 | -2.624709077 | -1.60602 |
| 28 | 8.334222 | -0.334222265 | -0.20451 |
| 29 | 6.52788 | 1.47211986 | 0.900768 |
| 30 | 3.818367 | -2.818366951 | -1.72452 |
| 31 | 7.431051 | -1.431051202 | -0.87564 |
| 32 | 7.431051 | 0.568948798 | 0.348131 |
| 33 | 12.85008 | 0.14992242 | 0.091735 |
| 34 | 8.334222 | -1.334222265 | -0.81639 |
| 35 | 5.624709 | -1.624709077 | -0.99413 |
| 36 | 9.237393 | 1.762606672 | 1.078512 |
| 37 | 10.14056 | -0.140564391 | -0.08601 |
| 38 | 12.85008 | 0.14992242 | 0.091735 |
| 39 | 6.52788 | 1.47211986 | 0.900768 |
| 40 | 8.334222 | -1.334222265 | -0.81639 |
| 41 | 7.431051 | -1.431051202 | -0.87564 |
| 42 | 9.237393 | -1.237393328 | -0.75714 |
| 43 | 7.431051 | -0.431051202 | -0.26375 |
| 44 | 10.14056 | 1.859435609 | 1.13776 |
| 45 | 5.624709 | -0.624709077 | -0.38225 |
| 46 | 9.237393 | -0.237393328 | -0.14526 |
| 47 | 13.75325 | 1.246751358 | 0.762868 |
| 48 | 6.52788 | -0.52788014 | -0.323 |
| 49 | 7.431051 | -0.431051202 | -0.26375 |
| 50 | 9.237393 | 0.762606672 | 0.466627 |
| 51 | 6.52788 | -0.52788014 | -0.323 |
| 52 | 8.334222 | 3.665777735 | 2.243033 |
| 53 | 11.04374 | -1.043735454 | -0.63865 |
| 54 | 8.334222 | -0.334222265 | -0.20451 |
| 55 | 4.721538 | -0.721538014 | -0.4415 |

PROBABILITY OUTPUT

| *Percentile* | *Y* |
|---|---|
| 0.909090909 | 1 |
| 2.727272727 | 3 |
| 4.545454545 | 3 |
| 6.363636364 | 3 |
| 8.181818182 | 3 |
| 10 | 3 |
| 11.81818182 | 4 |
| 13.63636364 | 4 |
| 15.45454545 | 4 |
| 17.27272727 | 4 |
| 19.09090909 | 4 |
| 20.90909091 | 4 |
| 22.72727273 | 4 |
| 24.54545455 | 5 |
| 26.36363636 | 5 |
| 28.18181818 | 5 |
| 30 | 5 |
| 31.81818182 | 5 |
| 33.63636364 | 6 |
| 35.45454545 | 6 |
| 37.27272727 | 6 |
| 39.09090909 | 6 |
| 40.90909091 | 6 |
| 42.72727273 | 6 |
| 44.54545455 | 7 |
| 46.36363636 | 7 |
| 48.18181818 | 7 |
| 50 | 7 |
| 51.81818182 | 7 |
| 53.63636364 | 7 |
| 55.45454545 | 7 |
| 57.27272727 | 7 |
| 59.09090909 | 7 |
| 60.90909091 | 7 |
| 62.72727273 | 7 |
| 64.54545455 | 8 |
| 66.36363636 | 8 |
| 68.18181818 | 8 |
| 70 | 8 |
| 71.81818182 | 8 |
| 73.63636364 | 8 |
| 75.45454545 | 8 |
| 77.27272727 | 8 |
| 79.09090909 | 8 |
| 80.90909091 | 9 |
| 82.72727273 | 9 |
| 84.54545455 | 10 |
| 86.36363636 | 10 |
| 88.18181818 | 10 |
| 90 | 11 |
| 91.81818182 | 12 |
| 93.63636364 | 12 |
| 95.45454545 | 13 |
| 97.27272727 | 13 |
| 99.09090909 | 15 |

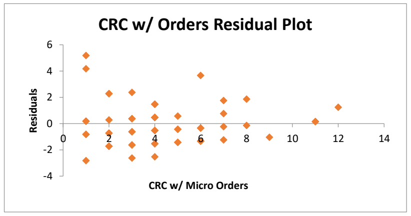


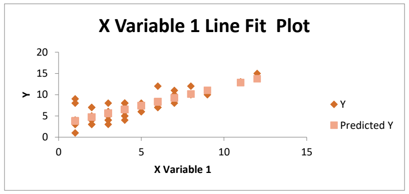


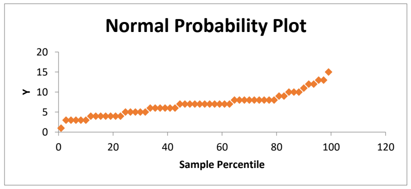
