## Supplemental Tale 3 for "Analysis of the Clinical Incidence and Correlation between Colorectal Cancer and Microorganisms"

SUMMARY OUTPUT

| *Regression Statistics* | |
|---|---|
| Multiple R | 0.605074 |
| R Square | 0.366115 |
| Adjusted R Square | 0.353437 |
| Standard Error | 1.162915 |
| Observations | 52 |

ANOVA

| | *df* | *SS* | *MS* | *F* | *Significance F* |
|---|---|---|---|---|---|
| Regression | 1 | 39.05456972 | 39.05457 | 28.87861 | 2.01971E-06 |
| Residual | 50 | 67.6185072 | 1.35237 | | |
| Total | 51 | 106.6730769 | | | |

| | *Coefficients* | *Standard Error* | *t Stat* | *P-value* | *Lower 95%* | *Upper 95%* | *ower 95.0%* | *Jpper 95.0%* |
|---|---|---|---|---|---|---|---|---|
| Intercept | 1.426015 | 0.288499323 | 4.942871 | 9.05E-06 | 0.846546896 | 2.005483 | 0.846547 | 2.005483 |
| X Variable 1 | 0.94151 | 0.175201135 | 5.373882 | 2.02E-06 | 0.589608422 | 1.293412 | 0.589608 | 1.293412 |

RESIDUAL OUTPUT

| *Observation* | *Predicted Y* | *Residuals* | *dard Residuals* |
|---|---|---|---|
| 1 | 2.367525 | 0.632474902 | 0.549282 |
| 2 | 3.309035 | -1.309035356 | -1.13685 |
| 3 | 2.367525 | -1.367525098 | -1.18765 |
| 4 | 2.367525 | 0.632474902 | 0.549282 |
| 5 | 1.426015 | 0.573985159 | 0.498486 |
| 6 | 2.367525 | -1.367525098 | -1.18765 |
| 7 | 2.367525 | -0.367525098 | -0.31918 |
| 8 | 3.309035 | -0.309035356 | -0.26839 |
| 9 | 1.426015 | -0.426014841 | -0.36998 |
| 10 | 2.367525 | -0.367525098 | -0.31918 |
| 11 | 2.367525 | -1.367525098 | -1.18765 |
| 12 | 3.309035 | -1.309035356 | -1.13685 |
| 13 | 2.367525 | -1.367525098 | -1.18765 |
| 14 | 1.426015 | -0.426014841 | -0.36998 |
| 15 | 2.367525 | -1.367525098 | -1.18765 |
| 16 | 3.309035 | -1.309035356 | -1.13685 |
| 17 | 1.426015 | -0.426014841 | -0.36998 |
| 18 | 3.309035 | -0.309035356 | -0.26839 |
| 19 | 2.367525 | 0.632474902 | 0.549282 |
| 20 | 1.426015 | -0.426014841 | -0.36998 |
| 21 | 2.367525 | 0.632474902 | 0.549282 |
| 22 | 2.367525 | 1.632474902 | 1.417747 |
| 23 | 2.367525 | -0.367525098 | -0.31918 |
| 24 | 2.367525 | -1.367525098 | -1.18765 |
| 25 | 2.367525 | 0.632474902 | 0.549282 |
| 26 | 1.426015 | -0.426014841 | -0.36998 |
| 27 | 4.250546 | -1.250545613 | -1.08606 |
| 28 | 2.367525 | 0.632474902 | 0.549282 |
| 29 | 2.367525 | -0.367525098 | -0.31918 |
| 30 | 1.426015 | 1.573985159 | 1.366951 |
| 31 | 3.309035 | 1.690964644 | 1.468544 |
| 32 | 2.367525 | -0.367525098 | -0.31918 |
| 33 | 2.367525 | 0.632474902 | 0.549282 |
| 34 | 3.309035 | 1.690964644 | 1.468544 |
| 35 | 2.367525 | 2.632474902 | 2.286212 |
| 36 | 3.309035 | 0.690964644 | 0.600079 |
| 37 | 4.250546 | -1.250545613 | -1.08606 |
| 38 | 3.309035 | -0.309035356 | -0.26839 |
| 39 | 3.309035 | -0.309035356 | -0.26839 |
| 40 | 3.309035 | -0.309035356 | -0.26839 |
| 41 | 4.250546 | -0.250545613 | -0.21759 |
| 42 | 2.367525 | -0.367525098 | -0.31918 |
| 43 | 2.367525 | -0.367525098 | -0.31918 |
| 44 | 4.250546 | 3.749454387 | 3.25627 |
| 45 | 3.309035 | -0.309035356 | -0.26839 |
| 46 | 2.367525 | 0.632474902 | 0.549282 |
| 47 | 3.309035 | 0.690964644 | 0.600079 |
| 48 | 4.250546 | -0.250545613 | -0.21759 |
| 49 | 1.426015 | 2.573985159 | 2.235416 |
| 50 | 5.192056 | 0.807944129 | 0.701671 |
| 51 | 3.309035 | -0.309035356 | -0.26839 |
| 52 | 2.367525 | -1.367525098 | -1.18765 |

PROBABILITY OUTPUT

| *Percentile* | *Y* |
|---|---|
| 0.961538462 | 1 |
| 2.884615385 | 1 |
| 4.807692308 | 1 |
| 6.730769231 | 1 |
| 8.653846154 | 1 |
| 10.57692308 | 1 |
| 12.5 | 1 |
| 14.42307692 | 1 |
| 16.34615385 | 1 |
| 18.26923077 | 1 |
| 20.19230769 | 1 |
| 22.11538462 | 1 |
| 24.03846154 | 2 |
| 25.96153846 | 2 |
| 27.88461538 | 2 |
| 29.80769231 | 2 |
| 31.73076923 | 2 |
| 33.65384615 | 2 |
| 35.57692308 | 2 |
| 37.5 | 2 |
| 39.42307692 | 2 |
| 41.34615385 | 2 |
| 43.26923077 | 2 |
| 45.19230769 | 3 |
| 47.11538462 | 3 |
| 49.03846154 | 3 |
| 50.96153846 | 3 |
| 52.88461538 | 3 |
| 54.80769231 | 3 |
| 56.73076923 | 3 |
| 58.65384615 | 3 |
| 60.57692308 | 3 |
| 62.5 | 3 |
| 64.42307692 | 3 |
| 66.34615385 | 3 |
| 68.26923077 | 3 |
| 70.19230769 | 3 |
| 72.11538462 | 3 |
| 74.03846154 | 3 |
| 75.96153846 | 3 |
| 77.88461538 | 3 |
| 79.80769231 | 4 |
| 81.73076923 | 4 |
| 83.65384615 | 4 |
| 85.57692308 | 4 |
| 87.5 | 4 |
| 89.42307692 | 4 |
| 91.34615385 | 5 |
| 93.26923077 | 5 |
| 95.19230769 | 5 |
| 97.11538462 | 6 |
| 99.03846154 | 8 |

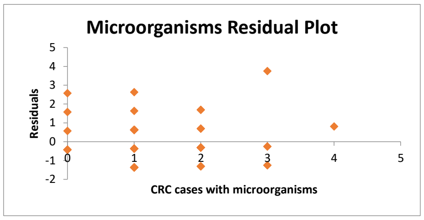


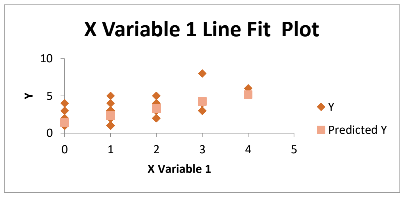


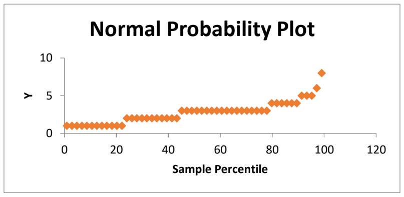
