## Supplemental Table 1 for "Analysis of the Clinical Incidence and Correlation between Colorectal Cancer and Microorganisms"

| CRC Case | Sex | Order/Test | BMI | Histological Type | Location | Tumor Size (cm) | pTNM | Differentiation | Co-occuring GI Disorder |
|---|---|---|---|---|---|---|---|---|---|
| **1** | male | Expedited covid 19, CORONAVIRUS (S | 21.1 | adenocarcinoma | left | | pT4 | | |
| **2** | female | two pre procedure and pre operative | 30.6 | adenocarcinoma | left | 0.7 | pT1 | well differentiated | |
| **3** | male | two pre procedure and pre operative | 22.1 | adenocarcinoma | left | | pT4 | moderately differentiated | |
| **4** | female | expedited covid19 | 22.38 | intramucosal adenocarcinoma | right | | | | GERD |
| **5** | female | coronavirus (sars cov 2 access labs), p | 21.5 | adenocarcinoma | right | 1 | pT3 | moderately differentiated | GERD |
| **6** | female | covid-19 PCR nasopharynx (bio-ref), C | 30.12 | adenocarcinoma | left | 2.5 | pT2 | | Ulcerative Colitis |
| **9** | male | expedited covid19 | 26.96 | adenocarcinoma | right | 3.5 | pT3 | moderately differentiated | GERD |
| **10** | male | pre procedure and pre operative covid | 24.1 | mixed neuroendocrine carcinoma-ade | left | | | | |
| **11** | female | pre procedure and pre operative covid | 23.9 | adenocarcinoma | left | | pT1 | well differentiated | |
| **12** | male | pre procedure and pre operative covid | 26 | adenocarcinoma | left | 15 | pT4 | moderately differentiated | |
| **13** | female | pre procedure and pre operative covid | 37.5 | adenocarcinoma | left | 0.4 | pT1 | moderately differentiated | GERD |
| **15** | male | 2019 coronavirus, coronavirus (sars co | 24.13 | adenocarcinoma | left | 3.5 | pT2 | moderately differentiated | GERD |
| **16** | female | two pre procedure and pre operative | 25.99 | adenocarcinoma | right | 4 | pT4 | moderately differentiated | |
| **17** | male | coronavirus (sars cov 2 access labs), p | 26.89 | adenocarcinoma | left | 1 | pT1 | moderately differentiated | GERD |
| **18** | male | coronavirus (sars cov 2 access labs), e | 28.12 | adenocarcinoma | left | | | moderately differentiated | |
| **19** | female | CCIRH rapid covid19, covid-19 PCR na | 25.6 | adenocarcinoma | left | 5.5 | pT3 | moderately differentiated | |
| **21** | female | four 2019 coronavirus, expedited cov | 24.2 | adenocarcinoma | left | 5.5 | pT3 | moderately differentiated | Type II Diabetes Mellitus |
| **23** | female | pre procedure and pre operative covid | 19.77 | signet-ring cell carcinoma | left | 3 | pT4 | poorly differentiated | |
| **24** | male | two 2019 coronavirus, pre procedure | 29.7 | adenocarcinoma | left | | | moderately differentiated | |
| **25** | female | two pre procedure and pre operative | 25 | adenocarcinoma | left | 4.3 | pT3 | moderately differentiated | |
| **26** | male | covid-19 PCR nasopharynx (BIOREF) | 28.94 | adenocarcinoma | left | 0.8 | | moderately differentiated | Type II Diabetes Mellitus |
| **27** | male | pre procedure and pre operative covid | 32.2 | adenocarcinoma | right | 4.2 | pT1 | well differentiated | GERD |
| **28** | female | expedited covid19 | 29.1 | mucinous adenocarcinoma | left | 1.8 | pT2 | | |
| **29** | male | pre procedure and pre operative covid | 36.78 | adenocarcinoma | right | 0.4 | pT3 | well differentiated | GERD |
| **30** | male | pre procedure and pre operative covid | 30.2 | adenocarcinoma | left | | | | GERD |
| **31** | female | pre procedure and pre operative covid | 31.16 | adenocarcinoma | right | 4.5 | pT2 | moderately differentiated | |
| **32** | female | pre procedure and pre operative covid | 23.6 | adenocarcinoma | left | | | moderately differentiated | |
| **33** | female | three pre procedure and pre operative | 19.1 | squamous cell carcinoma | left | 3.4 | pT2 | moderately differentiated | |
| **34** | male | pre procedure and pre operative covid | 25.1 | squamous cell carcinoma | left | 0.4 | | moderately differentiated | |
| **35** | female | expedited covid19 | 23.68 | neuroendocrine carcinoma | right | 6 | pT3 | poorly differentiated | |
| **36** | female | expedited covid | 28.3 | adenocarcinoma | right | 1.5 | pT1 | moderately differentiated | |
| **37** | female | coronavirus (sars cov 2 access labs) | 22.3 | adenocarcinoma | right | 4.5 | pT4 | moderately differentiated | |
| **38** | female | covid19 PCR nasopharynx (BIOREF) | 23.6 | adenocarcinoma | left | 5.5 | pT3 | well differentiated | |
| **39** | female | expedited covid19, coronavirus (sars | 30.8 | adenocarcinoma | left | 3 | pT2 | well differentiated | Ulcerative Colitis |
| **40** | male | two coronavirus (sars cov 2 access lab | 28 | mucinous adenocarcinoma | left | 4.5 | pT3 | | GERD |
| **41** | male | three coronavirus (sars cov 2 access la | 30.4 | adenocarcinoma | left | 0.3 | pT3 | moderately differentiated | Type II Diabetes Mellitus |
| **42** | female | coronavirus (sars cov 2 access labs), p | 33.2 | adenocarcinoma | left | 0.7 | pT2 | moderately differentiated | |
| **43** | male | pre procedure and pre operative covid | 27.1 | squamous cell carcinoma | left | 3 | pT2 | moderately differentiated | GERD |
| **45** | male | expedited covid19 | 23.7 | adenocarcinoma | left | 5.5 | pT3 | moderately differentiated | GERD |
| **46** | female | expedited covid19 | 24.24 | mucinous adenocarcinoma | right | 5 | pT3 | poorly differentiated | GERD |
| **47** | female | pre procedure and pre operative covid | 28.91 | adenocarcinoma | right | 4.5 | pT3 | moderately differentiated | |
| **48** | male | expedited covid19 | 26.1 | adenocarcinoma | right | 1.5 | pT2 | moderately differentiated | |
| **49** | male | pre procedure and pre operative covid | 26.2 | adenocarcinoma | left | 3 | pT3 | moderately differentiated | |
| **55** | male | coronavirus (sars cov 2 access labs), sa | 24.5 | adenocarcinoma | left | 3.5 | pT3 | moderately differentiated | Type II Diabetes Mellitus |
| **56** | male | 2019 coronavirus, two coronavirus (s | 30.4 | adenocarcinoma | right | 4 | pT2 | moderately differentiated | GERD |
| **57** | male | sars cov 2 rapid result antigen test | 26.8 | adenocarcinoma | left | 2.5 | pT1 | moderately differentiated | |
| **58** | male | expedited covid19 | 27.12 | adenocarcinoma | left | 3 | pT4 | moderately differentiated | GERD |
| **59** | male | four sars-cov-2 molecular POCT covid | 28.3 | adenocarcinoma | left | 4 | pT3 | poorly differentiated | |
| **60** | male | expedited covid19, two sars-cov-2 PC | 28.9 | adenocarcinoma | left | 4 | pT3 | moderately differentiated | Type II Diabetes Mellitus |
| **61** | male | pre procedure and pre operative covid | 30.4 | adenocarcinoma | left | 4.5 | pT2 | moderately differentiated | GERD |
| **62** | female | two sars-cov-2 RNA pre procedure an | 21.14 | adenocarcinoma | left | 2.5 | pT3 | moderately differentiated | |
| **64** | female | 2019 coronavirus, three pre procedur | 28.5 | adenocarcinoma | left | 4.5 | pT3 | poorly differentiated | |
| **66** | male | two sars-cov-2 RNA, expedited covid1 | 38.2 | adenocarcinoma | left | 5 | pT3 | moderately differentiated | Type II Diabetes Mellitus |
| **68** | male | expedited covid19, pre procedure anc | 27.4 | adenocarcinoma | left | 3.5 | pT3 | moderately differentiated | GERD |
| **69** | male | expedited covid19, 2019 coronavirus | 42.8 | mucinous adenocarcinoma | right | 5 | pT3 | moderately differentiated | |
| **73** | female | pre procedure and pre operative covid | 48 | adenocarcinoma | left | 3 | pT2 | moderately differentiated | |
| **75** | male | 2019 coronavirus, pre procedure and | 20.5 | adenocarcinoma | right | 7 | pT4 | well differentiated | |
| **76** | male | 2019 coronavirus | 27.83 | adenocarcinoma | right | 7.5 | pT3 | moderately differentiated | GERD |
| **77** | male | pre procedure and pre operative covid | 24.65 | adenocarcinoma | left | 7 | pT4 | | |
| **79** | male | sars-cov-2 PCR | 21.7 | adenocarcinoma | left | 3.2 | pT2 | moderately differentiated | |
| **80** | female | 2019 coronavirus, coronavirus (sars co | 27.21 | adenocarcinoma | left | 2 | pT2 | moderately differentiated | GERD |
| **81** | female | sars-cov-2 RNA, expedited covid19, c | 24.55 | adenocarcinoma | left | | | | GERD |
| **84** | female | coronavirus (sars cov 2 access labs) | 21.1 | adenocarcinoma | left | 7 | pT4 | poorly differentiated | |
| **85** | male | pre procedure and pre operative covid | 30.5 | adenocarcinoma | left | 4.5 | pT2 | moderately differentiated | |
| **93** | female | 2019 coronavirus, pre procedure and | 20.98 | adenocarcinoma | left | 6 | pT4 | moderately differentiated | GERD |
| **103** | female | pre procedure and pre operative covid | 19.9 | adenocarcinoma | left | 1.8 | pT2 | moderately differentiated | |
| **113** | male | expedited covid19, two sars cov 2 PCF | 28.1 | adenocarcinoma | right | 8.5,2 | pT3,pT1 | moderately differentiated, well differe | Type II Diabetes Mellitus & GERD |
| **114** | male | expedited covid + flu A/B, four pre pro | 31.74 | adenocarcinoma | right | 3.8 | pT3 | moderately differentiated | GERD |
| **115** | male | pre procedure and pre operative covid | 27.67 | adenocarcinoma | left | 3 | pT2 | poorly differentiated | |
| **117** | male | pre procedure and pre operative covid | 31.31 | adenocarcinoma | right | 4.5 | pT4 | moderately differentiated | GERD |
| **119** | male | expedited covid, expedited covid + flu | 29.35 | adenocarcinoma | left | 2.5 | pT2 | poorly differentiated | |
| **121** | female | expedited covid19 | 24.4 | adenocarcinoma | right | 4.6 | pT3 | moderately differentiated | |
| **124** | male | two expedited covid19, | 26.23 | medullary carcinoma | right | 11 | pT3 | | |
| **126** | male | 2019 coronavirus, pre operative and p | 30 | adenocarcinoma | left | 3.5 | pT3 | poorly differentiated | type II Diabetes Mellitus & GERD |
| **130** | male | two expedited covid, | 23 | adenocarcinoma | right | 10 | pT3 | moderately differentiated | |
| **136** | female | two expedited covid | 18.11 | adenocarcinoma | left | 2 | pT2 | moderately differentiated | type II Diabetes Mellitus |
| **137** | female | pre procedure and pre operative covid | 21.8 | adenocarcinoma | right | 1.5 | pT1 | moderately differentiated | |
| **142** | female | expedited covid19 | 24.8 | adenocarcinoma | right | 2.8 | pT3 | moderately differentiated | |
| **146** | male | three H&E/IHC stains, coronavirus (sa | 28.3 | adenocarcinoma | right | 3.5 | pT3 | moderately differentiated | |
| **149** | male | expedited covid19 | 31.34 | adenocarcinoma | left | 2.5 | | poorly differentiated | GERD |
| **152** | male | two expedited covid19 | 23.47 | adenocarcinoma | left | 2 | pT3 | | GERD |
| **153** | male | two 2019 coronavirus, covid19 PCR na | 27.67 | adenocarcinoma | left | 3.5 | pT1 | moderately differentiated | |
| **161** | female | two coronavirus (sars cov 2 access lab | 32.65 | adenocarcinoma | left | 2.5 | pT2 | moderately differentiated | GERD |
| **164** | female | coronavirus (sars cov 2 access labs), tv | 24.8 | adenocarcinoma | left | 2.5 | pT2 | moderately differentiated | GERD |
| **172** | male | pre procedure and pre operative covid | 29.07 | adenocarcinoma | right | 2.5 | pT4 | | |
| **178** | male | two expedited covid19, two pre proce | 26.05 | adenocarcinoma | right | 6.2 | pT4 | moderately differentiated | type II Diabetes Mellitus |
| **179** | female | expedited covid19, pre procedure anc | 26.15 | adenocarcinoma | left | 4 | pT4 | moderately differentiated | GERD |
| **181** | male | sars-cov-2 | 26.62 | adenocarcinoma | left | 1 | pT2 | moderately differentiated | type II Diabetes Mellitus |
| **182** | female | expedited covid19, Sars-Cov-2 | 30.73 | squamous cell carcinoma | left | 2 | | poorly differentiated | |
| **187** | male | coronavirus (sars cov 2 access labs), e: | 24.74 | adenocarcinoma | left | 4.5 | pT2 | moderately differentiated | GERD |
| **195** | male | expedited covid19, two covid19 PCR, | 18.19 | adenocarcinoma | left | 1.6 | pT4 | | GERD |
| **205** | male | two expedited covid, two coronavirus | 34.67 | adenocarcinoma | right | 0.3 | | moderately differentiated | |
| **206** | male | 2019 coronavirus, expedited covid19 | 30.06 | squamous cell carcinoma | left | 10.2 | pT3 | moderately differentiated | GERD |
| **207** | female | two CCIRH rapid covid | 25.9 | adenocarcinoma | left | 4.1 | pT1 | moderately differentiated | GERD |
| **208** | female | expedited covid, pre procedure and p | 17.12 | adenocarcinoma | left | 5 | pT2 | | GERD |
| **211** | male | coronavirus (sars cov 2 access lab) pre | 21.8 | adenocarcinoma | left | | | moderately differentiated | |
| **215** | male | three pre procedure and pre operative | 34.31 | adenocarcinoma | left | 6 | pT2 | moderately differentiated | |
| **217** | male | expedited covid19, two coronavirus (s | 22.78 | adenocarcinoma | left | 5 | pT4 | moderately differentiated | GERD |
| **219** | male | two expedited covid19, 2019 coronav | 30.73 | squamous cell carcinoma | left | 4 | pT4 | moderately differentiated | type II Diabetes Mellitus |
| **229** | male | expedited covid19, rapid SARS-COV-2 | 31.2 | adenocarcinoma | left | 4.5 | pT3 | moderately differentiated | |
| **239** | female | pre procedure and pre operative covid | 34.57 | adenocarcinoma | left | 0.1 | pT1 | well differentiated | |
